# Mapping AML heterogeneity – multi-cohort transcriptomic analysis identifies novel clusters and divergent ex-vivo drug responses

**DOI:** 10.1101/2023.03.29.23287896

**Authors:** Jeppe F Severens, E Onur Karakaslar, Bert A van der Reijden, Elena Sánchez-López, Redmar R van den Berg, Constantijn JM Halkes, Peter van Balen, Hendrik Veelken, Marcel JT Reinders, Marieke Griffioen, Erik B van den Akker

**Author notes:** Corresponding author: Erik B. van den Akker, PhD; Department of Biomedical Data Sciences, Leiden University Medical Center, Leiden, The Netherlands; Einthovenweg 20, 2333 ZC, Leiden, The Netherlands, 57. Disclosure of interest The authors declare no competing financial interests.

## Abstract

Subtyping of acute myeloid leukaemia (AML) is predominantly based on recurrent genetic abnormalities, but recent literature indicates that transcriptomic phenotyping holds immense potential to further refine AML classification. Here we integrated five AML transcriptomic datasets with corresponding genetic information to provide an overview (n=1224) of the transcriptomic AML landscape. Consensus clustering identified 17 robust patient clusters which improved identification of *CEBPA*-mutated patients with favourable outcomes, and uncovered transcriptomic subtypes for *KMT2A* rearrangements (2), *NPM1* mutations (5), and AML with myelodysplasia-related changes (AML-MRC) (5). Transcriptomic subtypes of *KMT2A*, *NPM1* and AML-MRC showed distinct mutational profiles, cell type differentiation arrests and immune properties, suggesting differences in underlying disease biology. Moreover, our transcriptomic clusters show differences in ex-vivo drug responses, even when corrected for differentiation arrest and superiorly capture differences in drug response compared to genetic classification. In conclusion, our findings underscore the importance of transcriptomics in AML subtyping and offer a basis for future research and personalised treatment strategies. Our transcriptomic compendium is publicly available and we supply an R package to project clusters to new transcriptomic studies.

## Introduction

In acute myeloid leukaemia (AML), recurrent genetic abnormalities (RGA) have been identified through systematic genomic studies.^1–5^ Based on these RGAs, the World Health Organization (WHO 2022) and International Consensus Classification (ICC 2022) define several AML subtypes, as well as a heterogeneous subtype of AML with myelodysplasia-related changes (AML-MRC).^6,7^ RGAs are essential for risk-stratification and are increasingly targeted with drugs.^8,9^

AML subclassification is genetics-based, but transcriptomics holds immense potential to refine AML classification further.^1–3,10–12^ Transcriptomic studies have led to the discovery of *CEBPA*-mutated AML^13,14^, and *NPM1*-mutated AML subtypes with different cell differentiation arrests and ex-vivo drug responses.^15,16^ Similar stratification would be beneficial for AML-MRC, given its heterogeneity.^17,18^ Still, a comprehensive examination of AML subtypes defined by gene expression has yet to be performed. Furthermore, the differentiation arrest state is known to modify drug response in AML^19^, and failing to account for this effect when comparing drug responses could skew conclusions.

Therefore, we integrated five mRNAseq datasets with corresponding genetic aberration data and annotated cases according to WHO and ICC 2022 standards. We outline AML’s transcriptomic landscape and define transcriptional subtypes with distinct gene expressions, genetic aberrations, and cell type arrests. We relate the clusters to ex-vivo drug responses independently of differentiation arrest and show how they superiorly capture differences in response compared to genetic classification. We provide all harmonised data and a transcriptional cluster predictor for future research. Our study underscores the importance of incorporating transcriptomic data in AML classification.

## Methods

### Transcriptomic data

We acquired transcriptomics data of primary AML patients from blood or bone marrow from BEAT^3,20^ (n = 425), TARGET^2^ (n = 145), TCGA^1^ (n = 150), and Leucegene^11,21–23^ (n = 399), and our in-house LUMC^24^ dataset (n = 95). Data statements and methods for transcriptome sequencing are available in the referenced studies.

We acquired quantified gene expression for BEAT, TARGET and TCGA from https://portal.gdc.cancer.gov/ (release 36) and implemented the same pipeline for Leucegene and LUMC to harmonise quantification. In short, FASTQ files were aligned and quantified with STAR^25^ to the GRCh38 reference genome^26^, using Gencode v36^27^ as the gene annotation index which included 60600 genes.

Gene expression count data were corrected with Combat-Seq^28^ for the variables “cohort”, “sex”, and “tissue”. We split Leucegene for the batch correction into Leucegene_stranded and Leucegene_unstranded, since different sequencing libraries were used. We removed 8057 genes that were not detected in all cohorts, leaving 52603 genes. Finally, we removed genes detected in less than 200 samples or with less than 300 counts leaving 41862 genes for our final dataset. We normalised the corrected count data using the geometric mean and variance stabilising transformation (VST)^29^ and quantified the remaining cohort-specific variation using kBET.^30^

### Genetic and patient data

We acquired genetic data for the samples from the referenced studies in the form of mutation and fusion calling, and cytogenetics data and clinical data on sex, age, blast percentage, and survival. Blacklisted fusions as reported by Arriba^31^ were removed from the fusion calling data.

We harmonised the data by standardising the annotation of gene names, fusion genes, and karyotyping. Using genetic data, we subclassified samples according to the WHO 2022^6^ and ICC 2022^7^. Samples for which we found no RGA and all genetic data available were annotated as “No RGA found”. We classified samples with missing data and “No RGA found” as “Inconclusive”.

### Clustering

We employed consensus clustering^32,33^ on the batch-adjusted gene expression. First, we created a weighted nearest neighbour graph^34^ using the 2000 most variable genes (MVGs). MVGs were selected via the median absolute deviation from samples with a blast percentage over 70% to minimise tumour microenvironment effects. Using the Leiden algorithm^35^ – with seed and resolution varied per iteration – we generated 300 cluster assignments from the graph for each n_clusters ranging from 10 to 20, totaling 3300 assignments.

From these 3300 assignments we created a consensus matrix with values ranging between 0 and 1 based on pairwise co-clustering. We then converted this matrix into a distance matrix (1 – consensus matrix) and conducted Ward.D2 hierarchical clustering. The final cluster count was determined based on the individual separation of WHO classes and clusters displaying differential traits.

### Cluster stability

To evaluate per sample clustering stability, we devised a stability score. We constructed a consensus matrix for each n_clusters (300 assignments) and subtracted each co-clustering value from 1 if it was below 0.5. Then, we took the mean of all values per sample as the stability score, which ranged from 0.5 to 1, with higher scores indicating less clustering ambiguity. To investigate correlation between cluster stability and blast percentage we performed a Spearman correlation test. Additionally, we generated tSNEs using 100 to 2500 MVGs to visually assess cluster stability.

### Cluster prediction

To predict cluster assignments we trained a one-vs-rest SVM per cluster. As input we used the uncorrected gene expression of the 2000 MVGs used for clustering. To select hyperparameters and evaluate performance we utilised 5×5 nested cross-fold validation.

To improve predictions we included a reject option using a minimum distance to the decision boundary. We determined this distance by looping over possible minimum values for the predictions of the inner fold. We selected the minimum value with the highest Kappa for the accepted inner fold samples and an accuracy < 0.5 for the rejected inner folds samples.

The final model was trained on the whole dataset, using 5-fold cross-validation to select hyperparameters and the minimum decision boundary distance.

### Differential gene expression analysis

Differential gene expression analysis between the clusters was performed using DESeq2^29^ using the corrected gene counts. We performed one-versus-rest analyses to identify differentially expressed genes in one cluster compared to all others. We annotated genes as transcription factors or coding for cell surface proteins using public databases.^36,37^

### Aberration enrichment analysis

To test if aberrations occurred more in a cluster than in others we first removed aberrations found in only one cohort or which occurred in less than 1% of the samples. We also included high *MECOM* expression in the analysis (VST expression > 6, based on the tail of a *MECOM* expression density plot). We tested for enrichment per aberration by performing an one-sided Fisher exact test for one cluster versus all others and adjusted p-values using the Benjamini-Hochberg (BH) procedure. We considered aberrations with a false discovery ratio (FDR) < 0.05 enriched.

### Survival analysis

We performed survival analysis using right censored overall survival data by generating Kaplan Meier (KM) curves on BEAT and TCGA survival data, comparing different groups of patients with the log rank test. We also performed Cox-regression using BEAT, TARGET and TCGA survival data for different patient groups and included cohort, sex and age as co-variables to analyse hazard ratios.

### Expression based score

We created cell type score to assess the differentiation arrest of AML samples, using the mean expression of 30 marker genes for six haematological cells.^38^ Additionally, we created immune phenotype scores for cytolytic infiltration and HLA I and HLA II antigen presenting cells using the mean expression of marker genes.^39^

### Drug response analysis

To analyse drug response differences, we used ex-vivo drug response data of 331 BEAT^3^ samples, quantified as area under the curve (AUC). We excluded drugs with less than 200 samples or missing data for any cluster, leaving 103 of the 123 drugs. We used a Kruskal-Wallis test for each drug with the AUC as response and clusters as groups to compare the average drug response per cluster. Drugs with a significant difference (FDR<.05) were analysed with one-sided Wilcoxon tests to identify clusters with low AUCs. Additionally, we performed a Kruskal-Wallis test for each drug with the ICC 2022 diagnosis as groups, to compare with clusters as groups.

Multivariate linear models (LM) were evaluated per drug to test if clusters were sensitive to a drug when adjusted for cell type, with AUC as response and cluster membership (one-versus-rest) and the six cell type scores as explaining variables. Similarly, we fitted LMs but with cluster membership and ICC 2022 diagnoses as variables. We considered clusters sensitive to a drug if the cluster membership’s FDR was below 0.05 and the LM coefficient was negative. All p-values were corrected using BH.

### Data Sharing Statement

The datasets generated and/or analysed during the current study are available from www.github.com/jeppeseverens/AMLmap.

### Code availability

All code used to generate results is available on reasonable request. The predictor is available from www.github.com/jeppeseverens/AMLmapR as an R package.

## Results

### Multi-cohort AML gene expression compendium

We collected 1224 RNAseq samples from adult (BEAT, TARGET, TCGA, Leucegene, LUMC) and paediatric (TARGET) cohorts with corresponding genetic and clinical data (**Figure 1A**). We quantified gene expression with the same pipeline and corrected counts for cohort, sex and source tissue (**Supplemental Figure 1**). Sample classification by their genetic data according to the WHO (**Figure 1C**) and ICC was successful for 97% of the samples. In line with previous reports, frequencies of the AML subtypes were similar for the adult cohorts but different between paediatric and adult cohorts (**Supplemental Table 1**), confirming that our dataset is representative of the AML landscape.^40,5^

**Figure 1:**
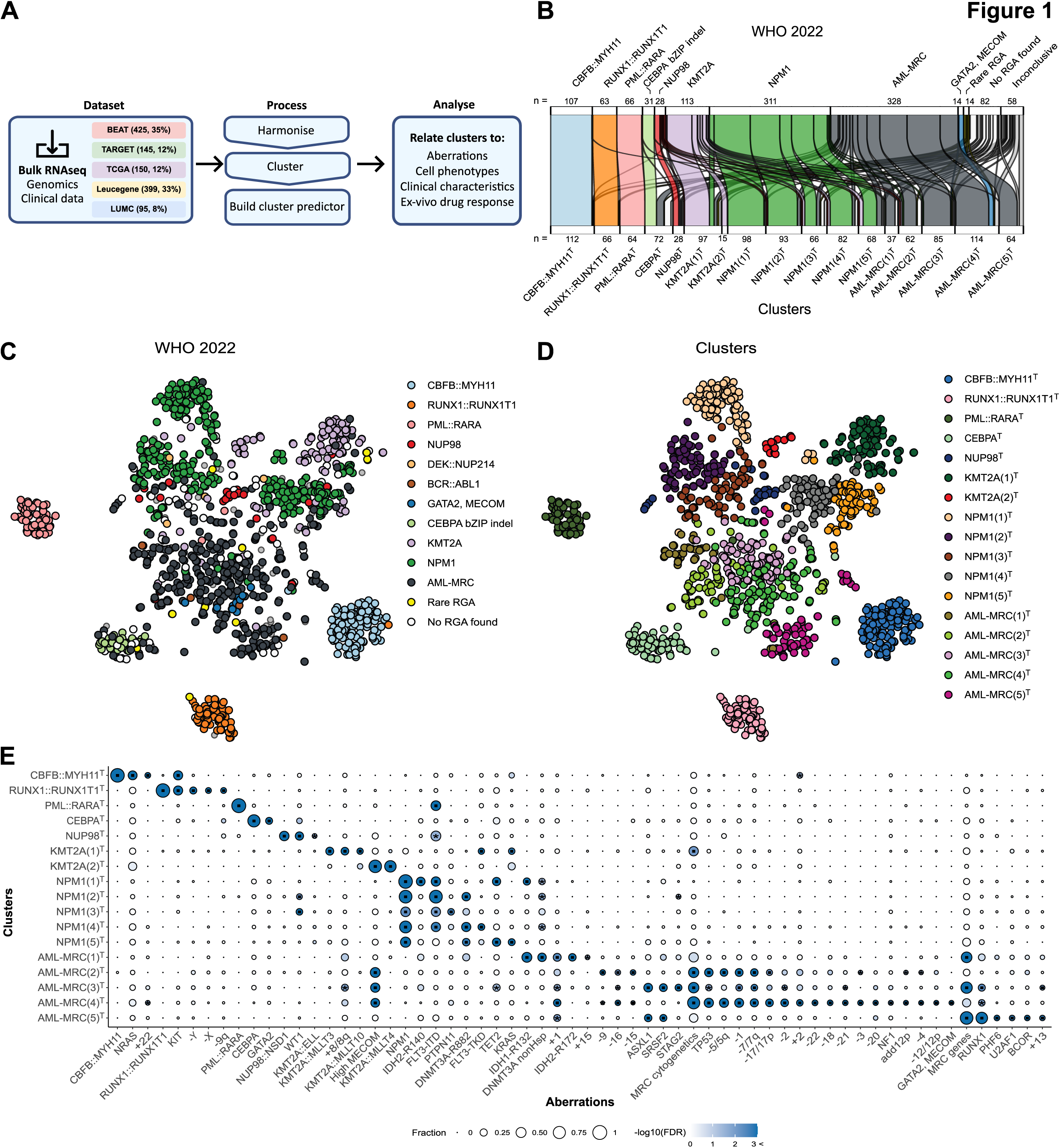
Transcriptomic analysis further stratifies AML. **A**) Flowchart of the used data and methods. **B)** Sankey plot showing the assignment of WHO 2022 diagnoses over the identified clusters. **C)** tSNE visualisation of the gene expression of patient samples. Each dot represents a patient. The samples are coloured according to the WHO 2022 subtyping of AML. **D)** The same tSNE visualisation as in **C**, but samples are coloured according to the 17 clusters. **E)** Dot plots that show enriched aberrations in the 17 clusters. The dots are coloured according to the adjusted p-value. The dots are sized according to the sample fraction with the aberration in the cluster. The x-axis shows the aberrations, and the y-axis shows the clusters. We only visualised enriched aberrations that occurred in at least 10% of the patients in a cluster.

### Transcriptomics define 17 AML clusters

Next, we assigned AML cases to 17 transcriptional clusters using consensus clustering (**Figure Supplemental Figures 2 & 3).** We named the clusters based on genetic diagnoses (**Figure 1B, Supplemental Figure 4 & 5**). As expected, the distribution over the clusters was different for paediatric and adult cohorts, exemplified by the large percentage of paediatric samples in the KMT2A^T^ clusters (26%), and adult samples in the NPM1^T^ (93%) and AML-MRC^T^ (94%) clusters (**Supplemental Table 1, Supplemental Figure 6**). However, samples of identical AML genetic subtypes from adult and paediatric cohorts clustered together, indicating that the 17 clusters capture differences in AML biology.

We examined clustering robustness using the stability score (**Supplemental Figure 7**). Median clustering stability was high (0.97-1.00), with AML-MRC(3)^T^ showing the lowest stability. A correlation test revealed a significant but weak correlation (rho = 0.18, p-value<.001) between blast percentage and clustering robustness, but blast percentage varied greatly in clusters. tSNEs generated using different MVGs (**Supplemental Figure 8**) were stable from 500 to 2500 features. These results show that clustering was robust and only weakly influenced by blast percentage.

We developed a transcriptional cluster predictor using uncorrected counts as input (accuracy = 0.90), demonstrating the persistence of expression patterns. The quality of the predictor was further improved (accuracy = 0.95) by including a reject option (10% rejected) (**Supplemental Figure 9**).

Next, we tested for enrichment of mutations, fusions and cytogenetic aberrations (n=102) (**Figure 1E**, p-values and frequencies in **Supplemental Table 2**). Four transcriptomic clusters corresponded to singular genetic AML subtypes: RUNX1::RUNX1T1^T^ (*RUNX1::RUNX1T1*: 94%, FDR<.001), CBFB::MYH11^T^ (*CBFB::MYH11*: 95%, FDR<.001), PML::RARA^T^ (*PML::RARA*: 100%, FDR<.001), and NUP98^T^ (*NUP98::NSD1*: 45%, FDR<.001). Risk-stratification for survival based on transcriptional subtypes performed similarly to genetics (**Supplemental Figure 10 & 11**). We identified no enrichment for *BCR::ABL1* and *DEK::NUP214*, possibly due to their limited occurrence. For *KMT2A* rearrangements, *CEBPA* mutations, *NPM1* mutations, and AML-MRC, we found evidence that transcriptomics can refine subtyping, as described below.

### Transcriptome analysis identifies two *KMT2A*-related clusters

The WHO classification defines a single *KMT2A*-rearranged subtype (*KMT2A*-r), while the ICC recognises *KMT2A::MLLT3* and other *KMT2A* fusions as distinct.^6,7^ We identified two *KMT2A* fusion clusters. KMT2A(1)^T^ was significantly enriched for *KMT2A::MLLT3* (31%, FDR<.001)*, KMT2A::MLLT10* (19%, FDR<.001) and any *KMT2A* fusion (67%, FDR<.001), while KMT2A(2)^T^ was enriched for *KMT2A::MLLT4* (67%, FDR<.001) and high *MECOM* expression (80%, FDR<.001) (**Figures 2A** **& B**). Interestingly, we found cases with *NPM1* mutations and trisomy 8/8q localised in KMT2A(1)^T^, indicating that these lead to *KMT2A* fusion-like gene expression.

**Figure 2:**
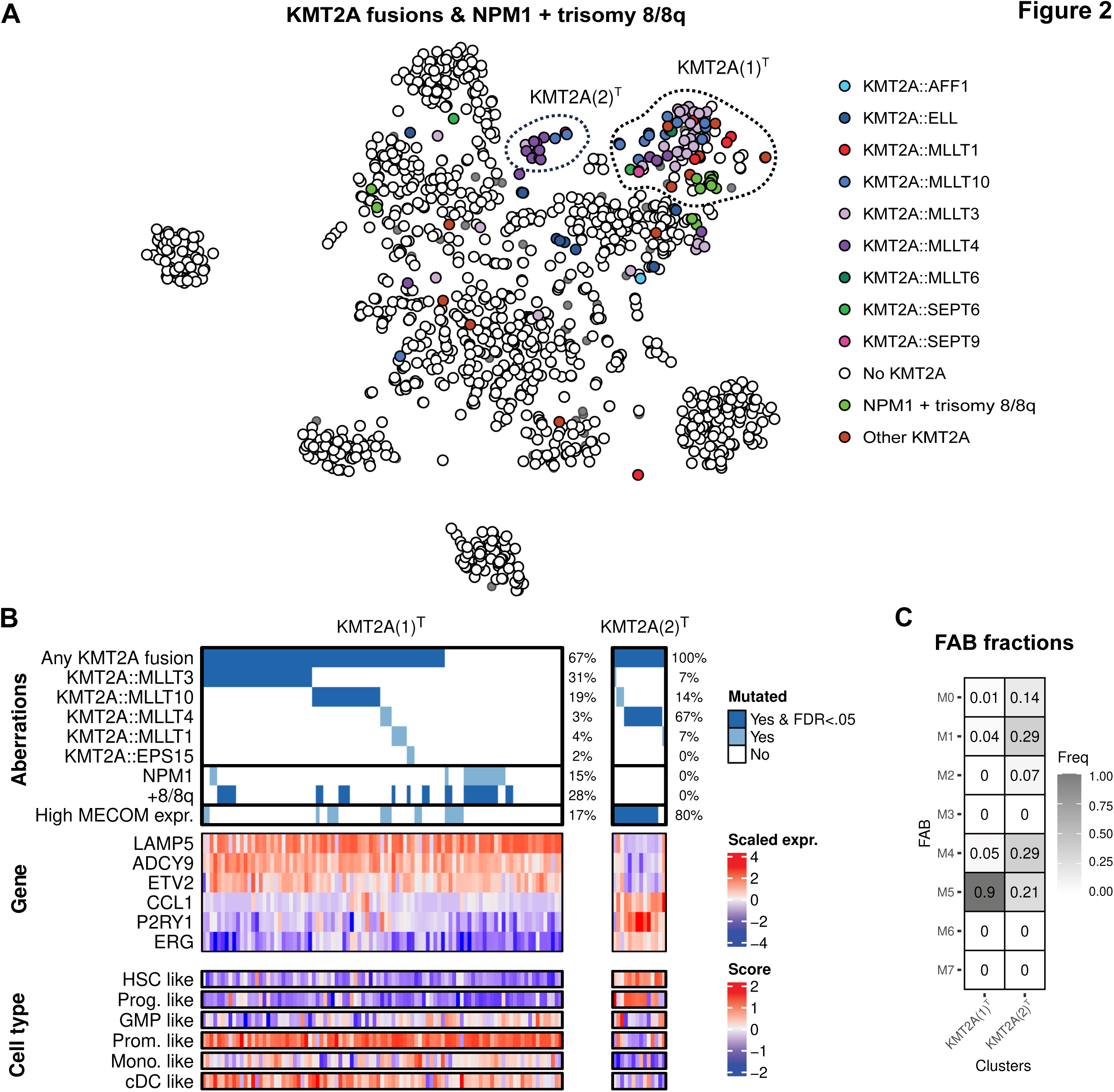
Transcriptome analysis identifies two *KMT2A*-related clusters. **A**) tSNE visualisation of patient samples, coloured according to *KMT2A*-fusion or *NPM1* mutation and trisomy 8. **B)** Waterfall plot of aberrations in the KMT2A clusters, including the percentage of samples with the aberration. The plot is combined with heatmaps showing the expression of marker genes and cell type scores. The columns are samples, which are split according to transcriptional clusters. **C)** Fraction of FAB annotations per cluster. HSC = hematopoietic stem cells, Prog. = progenitor, GMP = granulocyte-monocyte progenitor, Prom. = promonocytes, Mono. = monocytes, cDC = conventional dendritic cells

The genes *LAMP5*, and *ADCY9* showed high expression in KMT2A(1)^T^ and low expression in KMT2A(2)^T^ (**Figure 2B**), and all have been shown to contribute to AML pathogenesis^41,42^. Additionally, the transcription factor (TF) *ETV2* was highly expressed in KMT2A(1)^T^, while the TF *ERG* displayed high expression in KMT2A(2)^T^. The cell type scores revealed KMT2A(1)^T^ to have a significantly higher promonocyte-like score (FDR<.001), while KMT2A(2)^T^ was more hematopoietic stem cell (HSC)-like (FDR<.001) (**Figure 2B**, **Supplemental Figure 12**). FAB annotations showed similar results for KMT2A(1)^T^, which had a high M5 (monocytic leukaemia) fraction (90%), while KMT2A(2)^T^ was more mixed (**Figure 2C**). Overall, we found that gene expression-based separation of *KMT2A*-r did not align with the ICC 2022 classification.

### The CEBPA^T^ cluster indicates a favourable prognosis

As acknowledged in the ELN2022, patients with a *CEBPA* bZIP inframe mutation have a favourable prognosis.^8,43^ We identified a transcriptional CEBPA^T^ cluster significantly enriched for mutated *CEBPA* cases (72%, FDR<.001), with 42% of the samples having a *CEBPA* bZIP indel, either as single mutation or combined with an N-terminal frameshift mutation (**Figure 3A** **& B**). The remaining samples contained other mutations in the bZIP area or N-terminal region or had no detectable *CEBPA* mutation. Of note, a single *CEBPA* bZIP indel case resided outside the CEBPA^T^ cluster. This patient had an IDH-R132 mutation with a VAF=0.47, while the CEBPA bZIP in-frame mutation had a VAF=0.21. This finding suggests that the *IDH*-R132 mutation dominates the expression pattern, placing this case in cluster AML-MRC(1)^T^. Conversely, all CEBPA^T^ cluster patients showed similar favourable outcomes (log-rank test: p-value=.80), irrespective of whether the *CEBPA* bZIP inframe mutation was detected (**Figure 3C**). The CEBPA^T^ cluster thus marks patients with a favourable outcome regardless of *CEBPA* mutation detection, which the CEBPA^T^ expression profile can detect.

**Figure 3:**
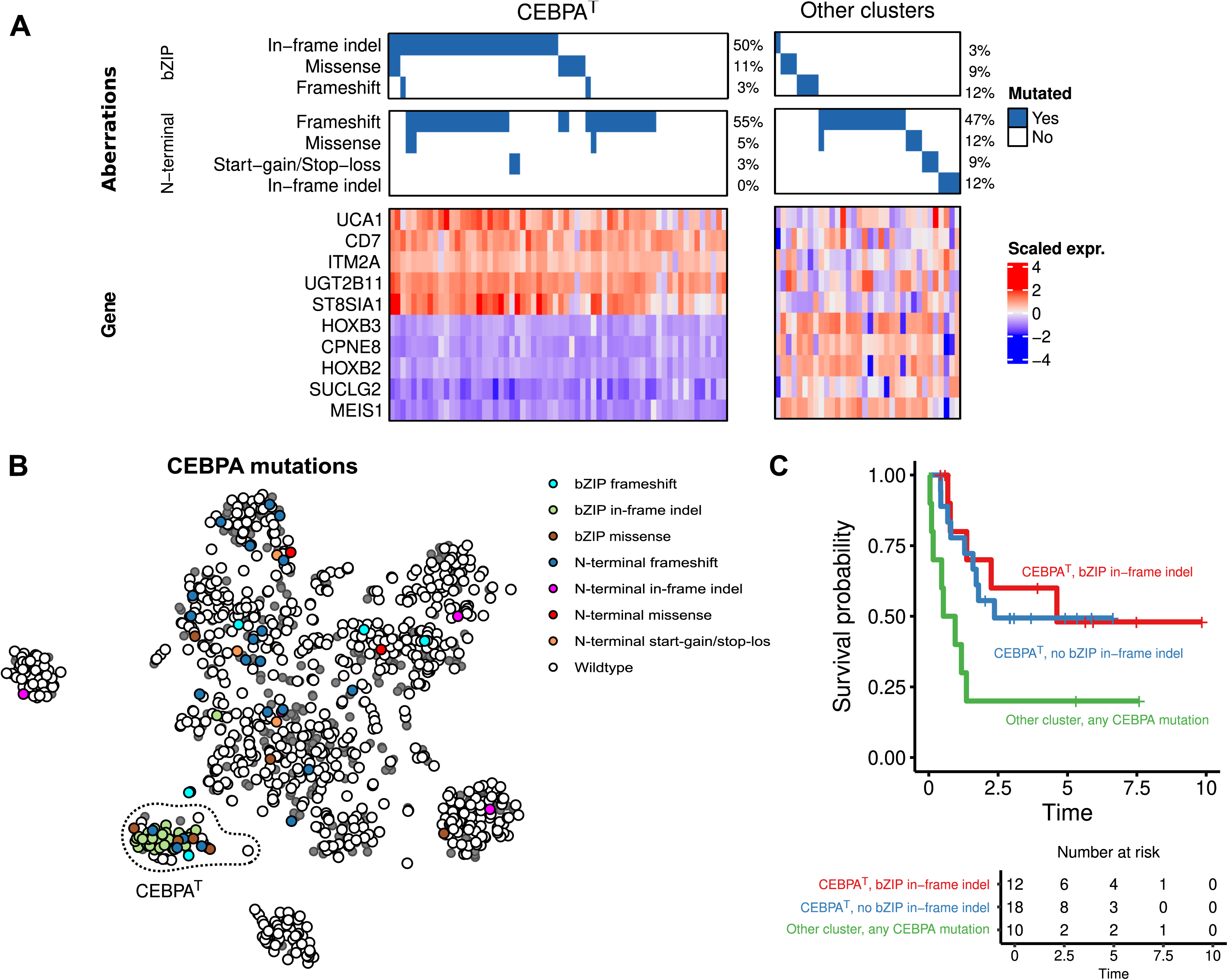
The CEBPA^T^ cluster indicates a favourable prognosis. **A**) Waterfall plot and gene expression heatmap of all samples in the CEBPA^T^ cluster and samples with a *CEBPA* mutation located outside the CEBPA^T^ cluster. **B)** tSNE visualisation of patient samples, coloured according to the type of *CEBPA* mutation. For samples with multiple *CEBPA* mutations, we used the ordering as in **A** to decide which mutation to display. **C)** Kaplan-Meier curve of the survival of BEAT and TCGA patients in and outside the CEBPA^T^ cluster.

### Gene expression profiling identifies five transcriptional *NPM1*-related clusters

The 2022 WHO and ICC classifications include one subtype of *NPM1*-mutated AML.^6,7^ However, we identified five clusters enriched for mutated *NPM1* (**Figure 4**). We observed elevated expression of *HOXA3*, *HOXB5*, and *MEIS1* (**Figure 4A**), which has been earlier associated with *NPM1* mutations.^44^ Interestingly, *NPM1* mutation-lacking samples generally also exhibited high expression of these genes, suggesting that there are alternative mishaps that disrupt these genes leading to *NPM1* mutated-like AML.

**Figure 4:**
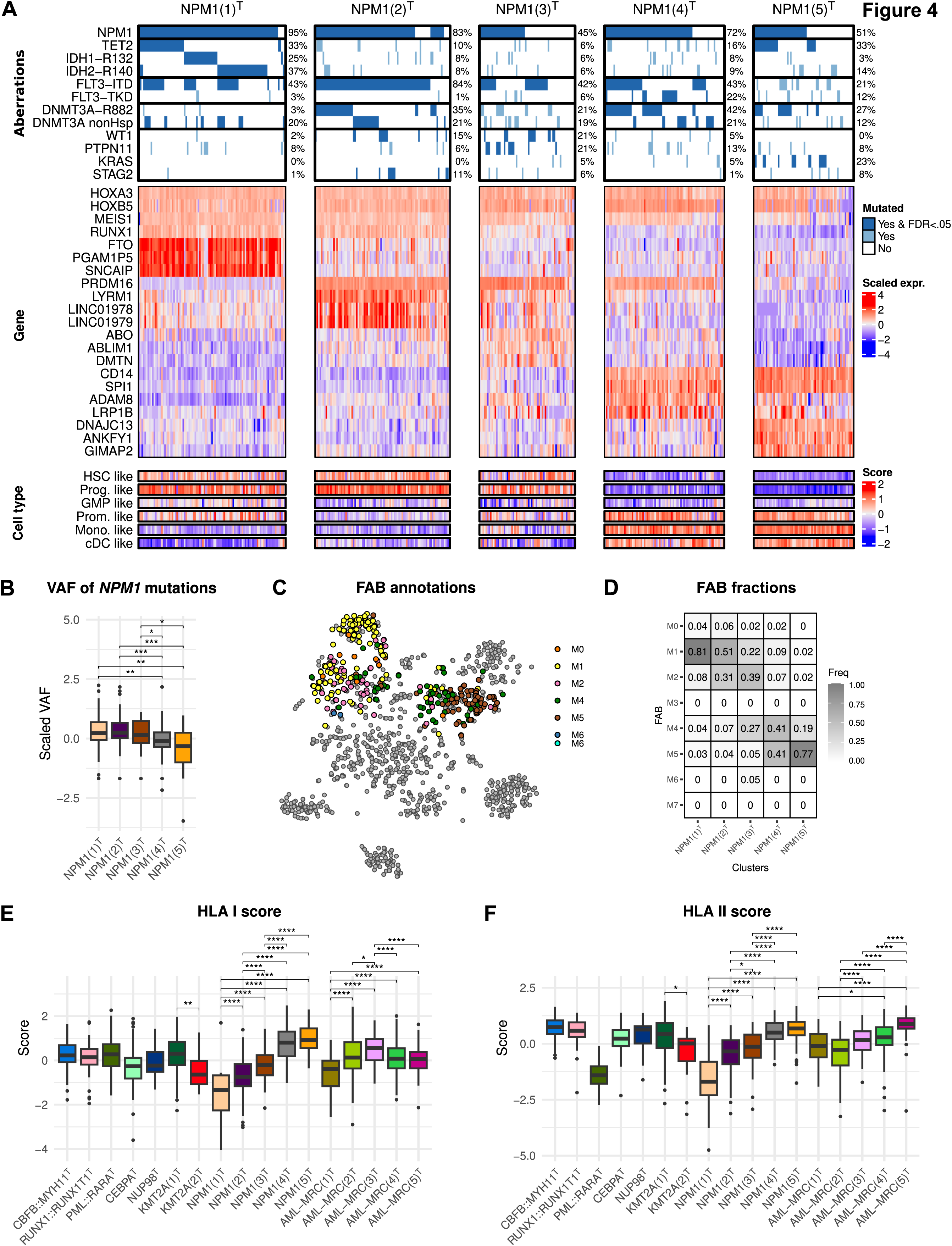
Gene expression profiling identifies five transcriptional *NPM1*-related clusters. **A**) Waterfall plot of aberrations in the *NPM1*-related clusters, including the percentage of samples with the aberration. The plot is combined with heatmaps showing the expression of marker genes and cell type scores. The columns are samples, which are split according to transcriptional clusters. **B)** Boxplot showing the scaled variant allele frequency (VAF) of mutated *NPM1* from the BEAT, Leucegene, and LUMC cohorts. The VAF was scaled per gene and study to allow for a combined analysis. We used a two-sided Wilcoxon test to test for statistical differences and Benjamini-Hochberg to adjust p-values for multiple testing. **C)** tSNE visualisation of patient samples, coloured according to the FAB annotation. Only *NPM1*-related clusters are coloured; the rest are in grey. **D)** Fraction of FAB annotations per cluster. **E)** and **F)** show boxplots of HLA I and HLA II antigen presenting cell scores, respectively. Tests were performed as in **B**, but were only done between the two KMT2A^T^, the five NPM1^T^ and the five AML-MRC^T^ clusters. FDR values: * < 0.05, ** < 0.01, *** < 0.005, **** < 0.001. HSC = hematopoietic stem cells, Prog. = progenitor, GMP = granulocyte-monocyte progenitor, Prom. = promonocytes, Mono. = monocytes, cDC = conventional dendritic cells, VAF = variant allele frequency

NPM1(1)^T^ exhibited the highest percentage (95%, FDR<.001) of *NPM1* mutated samples and was significantly enriched for *IDH1*-R132 (25%, FDR<.001), *IDH2*-R140 (37%, FDR<.001), and *TET2* (33%, FDR<.001) co-mutations (**Figure 4A**). NPM1(2)^T^ samples were enriched for *FLT3*-ITD mutations (84%, FDR<.001), but *FLT3*-ITD was also enriched in NPM1(1)^T^, NPM1(3)^T^ and NPM1(4)^T^ (42-43%, all FDR<.001). Additionally, NPM1(4)^T^ and NPM1(5)^T^ had a significantly lower variant allele frequency for mutated *NPM1* (**Figure 4B**). We found two *NPM1*::*MLF1* cases in our compendium, which both clustered in NPM1(3)^T^. *NPM1*::*MLF1* has been shown to localise in the cytoplasm^45^, like mutated *NPM1*, possibly leading to a similar expression profile as *NPM1*-mutated cases.

Each of the *NPM1*-related clusters exhibited unique marker genes (**Figure 4A**). For instance, *FTO* expression was high in NPM1(1)^T^. Additionally, *LYRM1*, *ADAM8*, and *DNAJC13* were elevated in NPM1(2)^T^, NPM1(4)^T^, and NPM1(5)^T^, respectively. NPM1(3)^T^ had a less distinct expression pattern, suggesting a more heterogeneous cluster. Also, we observed differential expression of TFs (*RUNX1*, *PRDM16*, *SPI1*)^46,47^ – even in samples lacking the *NPM1* mutation – and TF expression aligned with cell differentiation stages.

NPM1(1)^T^ and NPM1(2)^T^ displayed a HSC-like expression pattern, NPM1(3)^T^ was mixed, whereas NPM1(4)^T^ and NPM1(5)^T^ were more differentiated (**Figure 4A**, **Supplemental Figure 12**). FAB annotations showed additional differences, with NPM1(5)^T^ containing fewer M4 (myelomonocytic leukaemia) but more M5 (monocytic leukaemia) cases than NPM1(4)^T^ (**Figure 4C** **& D**). Using scores for HLA I and HLA II antigen-presenting cells^39^ we found NPM1(1)^T^ to have significantly lower HLA I (FDR<.001) and HLA II (FDR<.001) scores than the other clusters (**Figure 4E** **& F**). NPM1(1)^T^ and NPM1(5)^T^ patients were significantly older (FDR<.05), while NPM1(3)^T^ patients were younger (FDR<.01) (**Supplemental Figure 6F**). Our findings emphasise the existence of distinct *NPM1*-related subsets, highlighting the limitations of relying solely on genetic classification.

### Gene expression profiling identifies five transcriptional AML-MRC-related clusters

The ICC 2022 divides AML-MRC into three groups based on TP53 mutations, myelodysplasia-related gene mutations, and cytogenetic abnormalities.^7^ Our study identified five gene expression-based AML-MRC related clusters (**Figure 5**), with varying fractions of *TP53* mutations, MRC gene mutations, and cytogenetic abnormalities.

**Figure 5:**
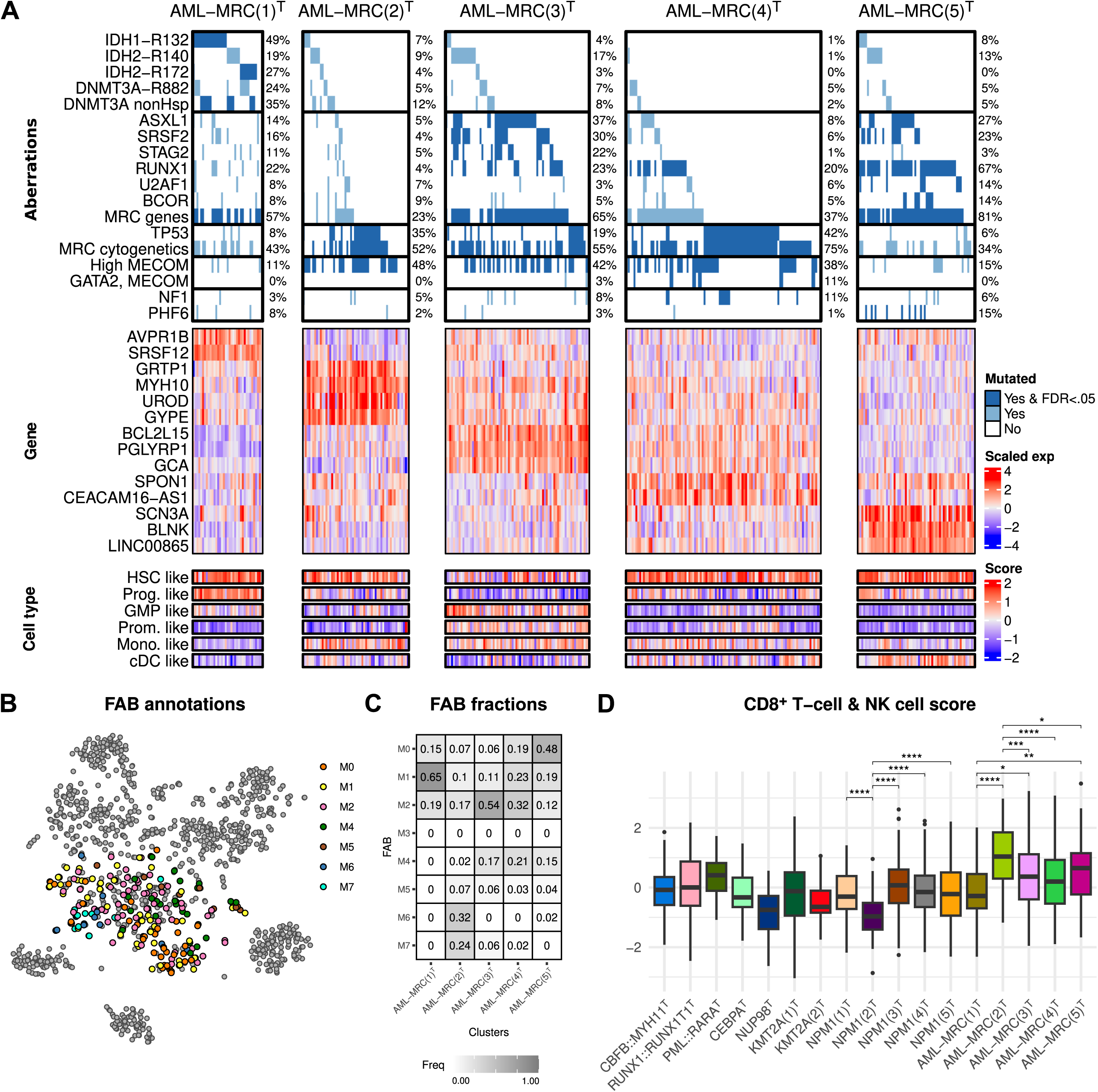
Gene expression profiling identifies five transcriptional AML-MRC-related clusters. **A**) Waterfall plot of aberrations in the AML-MRC-related clusters, including the percentage of samples with the aberration. We did not plot enriched individual large chromosomal mishaps. The plot is combined with heatmaps showing the expression of marker genes and cell type scores. The columns are samples, which are split according to transcriptional clusters. MRC genes are *ASXL1*, *BCOR*, *EZH2*, *RUNX1*, *SF3B1*, *SRSF2*, *STAG2*, *U2AF1*, or *ZRSR2*. Cytogenetic abnormalities are the ICC 2022 aberrations that define AML-MRC with cytogenetic abnormalities. **B)** tSNE visualisation of patient samples, coloured according to the FAB annotation. Only AML-MRC clusters samples are coloured are coloured; the rest are in grey. **C)** Fraction of FAB annotations per cluster. **D)** Boxplots of cytolytic cell score per cluster. We used a two-sided Wilcoxon test to test for statistical differences and Benjamini-Hochberg to adjust p-values for multiple testing. Tests were performed only between the two KMT2A^T^, the five NPM1^T^ and the five AML-MRC^T^ clusters. FDR values: * < 0.05, ** < 0.01, *** < 0.005, **** < 0.001. HSC = hematopoietic stem cells, Prog. = progenitor, GMP = granulocyte-monocyte progenitor, Prom. = promonocytes, Mono. = monocytes, cDC = conventional dendritic cells

Despite sharing these mishaps, each cluster had unique characteristics (**Figure 5A**). AML-MRC(1)^T^ was characterised by *IDH1*-R132 (49%, FDR<.001) and *IDH2*-R170 (27%, FDR<.001) mutations. A *DNMT3A* and *IDH1/2* mutated subtype has been reported^5^, but 41% of the AML-MRC(1)^T^ cases lacked *DNMT3A* mutations. AML-MRC(2)^T^, AML-MRC(3)^T^, and AML-MRC(4)^T^ were all enriched (FDR<.001) for *TP53* mutations, cytogenetic abnormalities and high *MECOM* expression, and AML-MRC(3)^T^ also contained a large fraction of mutated MRC genes (65%, FDR<0.001). AML-MRC(5)^T^ stood out with the highest fraction of mutated MRC genes cases (81%, FDR<.001) and the lowest fraction of *TP53* mutations (6%) and cytogenetic abnormalities (34%).

We found marker genes for all clusters (**Figure 5A**). For instance, high S*RSF12* marked AML-MRC(1)^T^, and *LINC00865* marked AML-MRC(5)^T^. AML-MRC(2)^T^ presented high glycophorin genes and *UROD* expression, suggesting an association with acute erythroid leukaemia.^48–50^ Distinct cell differentiation scores further highlighted differences (**Figure 5A-C, Supplemental Figure 12)**. For example, AML-MRC(1)^T^ showed high progenitor-like scores, with 65% M1 (minimal maturation) cases, and AML-MRC(3)^T^ showed a more differentiated pattern, with 54% M2 (significant maturation) cases. AML-MRC(2)^T^ was the only cluster with M6 (erythroid leukaemia) – in line with high expression of erythrocyte cell markers – and M7 (megakaryocytic leukaemia) cases. Additionally, high cytolytic cell infiltration has been reported for AML-MRC cases.^39^ Using the same score (**Figure 5D**) we found that the cytolytic infiltration was significantly (FDR<.05) lower for AML-MRC(1)^T^ and higher for AML-MRC(2)^T^ compared to the other AML-MRC clusters. Our results demonstrate that different AML-MRC transcriptomic clusters can be identified, showing genetic enrichments that do not necessarily align with the ICC 2022 classification.

### AML clusters exhibit cell type-independent differences in ex-vivo drug responses

Finally, we assessed the drug sensitivity of the transcriptional AML subtypes. Using ex-vivo drug response data, we discovered 101 drug-cluster combinations with significantly lower resistance (FDR<.05), of which 21 combinations remained statistically significant (FDR<.05) when adjusting for cell differentiation status (**Figure 6**, **Supplemental Figure 13, Supplemental Table 3**).

**Figure 6:**
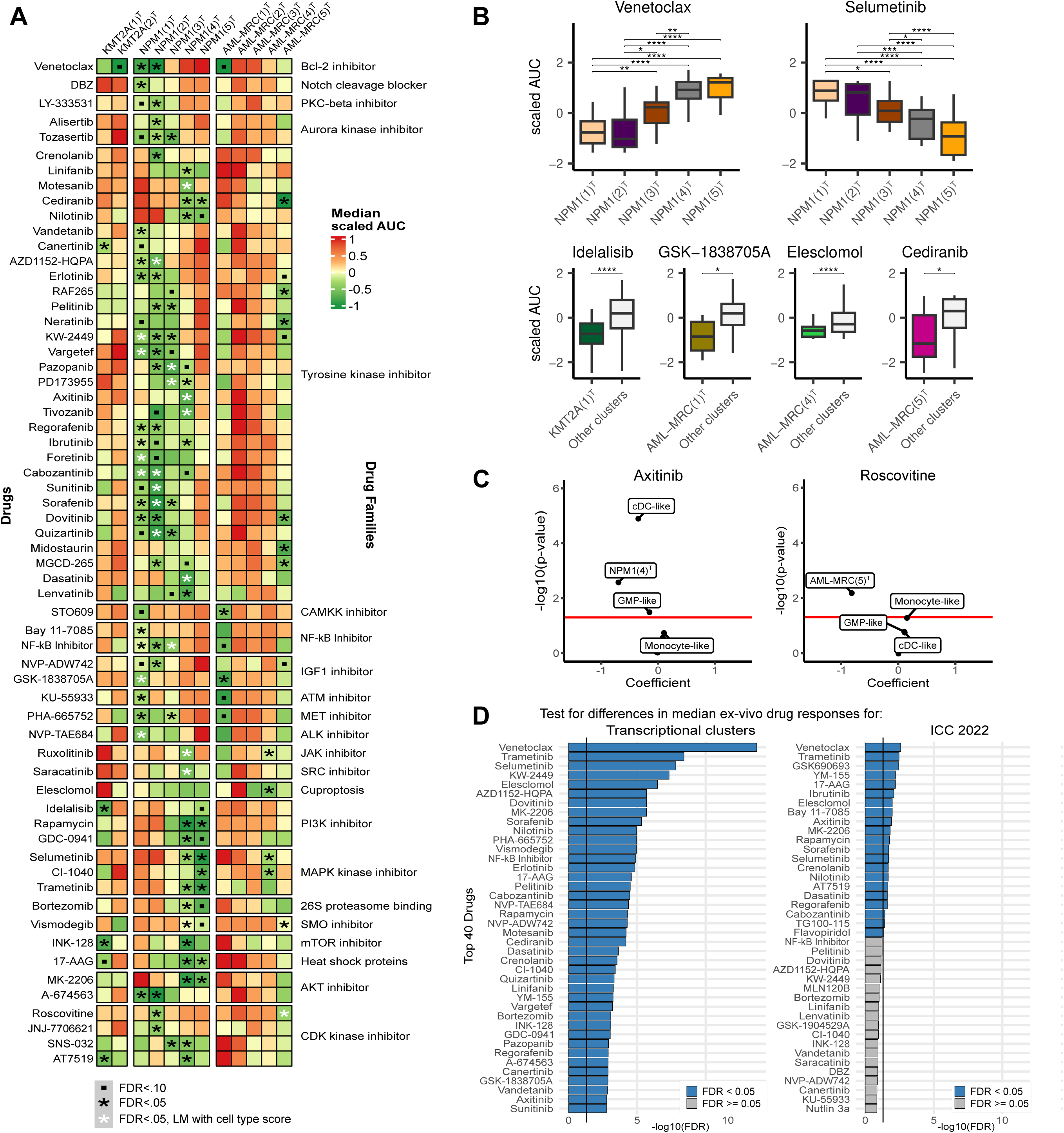
AML clusters exhibit cell type-independent differences in ex-vivo drug responses. **A**) Heatmap coloured according to the median scaled area under the curve (AUC) of the ex-vivo drug response per drug and cluster. On the left is the drug name, and on the right is the drug family. A green colour indicates a lower median AUC for the drug for the samples in the cluster compared to the other clusters, indicating a strong drug response. Red indicates a higher median AUC, meaning a weak drug response. **B)** Boxplots showing ex-vivo drug responses for a selection of drugs. We performed significance testing using a two-sided Wilcoxon test. FDR values: * < 0.05, ** < 0.01, *** < 0.05, **** < 0.001. **C)** Plots of the multivariate linear models with cluster membership and the six cell type scores as independent variables and AUC as the dependent variable. On the x-axis, the coefficient of the variables in the models is shown, and the y-axis shows the –log10 of the p-value for each variable. The shown p-values are not corrected and are for visualisation to indicate variable importance in the multivariate model. The corrected p-values of the cluster membership variable are shown in **Supplemental Table 3.** The red line indicates a p-value of 0.05. **D)** Barplots of the top 40 drugs with highest corrected p-values for Kruskal-Wallis tests between ex-vivo drug response and clusters or ICC 2022 diagnosis to test if there were significant differences in the median AUCs. All p-values were corrected with Benjamini-Hochberg.

The ex-vivo drug responses between *NPM1*-related clusters were often divergent, exemplified by venetoclax and selumetinib (**Figure 6A,B**). NPM1(1)^T^, NPM1(2)^T^ and NPM1(3)^T^ mostly responded positively to tyrosine kinase inhibitors and CDK kinase inhibitors. NPM1(4)^T^ and NPM1(5)^T^ samples were more sensitive to PI3K and MAPK kinase inhibitors. We also found drugs where only one cluster was responsive, exemplified by axitinib for NPM1(4)^T^ where this effect remained significant (FDR<.05) when controlled for cell type scores (**Figure 6C**).

Several drugs demonstrated favourable ex-vivo responses in KMT2A(1)^T^ compared to the other clusters, exemplified by idelalisib. For KMT2A(2)^T^ we found no significant responsive drugs, but testing was limited due to small cluster size. For the AML-MRC clusters, most drugs showed strong resistance. Still, specific drugs were more effective for AML-MRC(1)^T^, AML-MRC(4)^T^, and AML-MRC(5)^T^ (**Figure 6A, B**), suggesting potential for targeted treatments in this diverse, high-risk patient group.

Next, we examined if transcriptional clusters provide insights beyond genetic classifications. Comparing the AUCs of each drug between groups, we found 71 drugs with significantly different (FDR<.05) median AUCs between the clusters, while only 21 drugs were significantly different between ICC 2022 classes (**Figure 6D**). Additionally, 57 of the 101 cluster-drug combinations remained significant (FDR<.05) when cluster membership and ICC 2022 diagnosis were included in a LM, suggesting that the transcriptional clusters offer information beyond genetic classification.

Overall, our findings offer novel opportunities for targeted therapy in AML. We observed effective drug responses even after adjusting for differentiation status, possibly allowing gene expression-based subtypes to guide treatment strategies.

## Discussion

This study presents an overview of transcriptomics in AML and provides a framework for transcriptional subtyping. We integrate multiple cohorts to identify 17 robust transcriptional subtypes that subclassify ∼75% of our datasets’ patients. We make the harmonised data and a cluster predictor publicly available, facilitating future research.

For the CEBPA^T^ cluster, we show that patients without a detected *CEBPA* bZIP indel mutation still have similar favourable survival. Patients without the canonical *CEBPA* mutation in the CEBPA^T^ cluster could be explained by undetected mutations, given the complexities of CEBPA sequencing. Also, *CEBPA* hypermethylation has been described to lead to a similar expression profile.^51^ The use of the CEBPA^T^ gene signature for risk stratification could be a relevant alternative to detect these favourable-outcome patients.

KMT2A(1)^T^ mainly featured *KMT2A* with the fusion partners *MLLT3*, *MLLT10* and *MLLT1* – all TFs in the super elongation complex whose perturbation leads to disrupted hematopoietic lineage commitment.^52^ In contrast, KMT2A(2)^T^ featured *KMT2A::MLLT4*, which is thought to cause leukaemia by promoting self-association^53^. Interestingly, *MLLT3*, *MLLT10* and *MLLT1* all fuse a specific region of *KMT2A*, but *MLLT4* shows less specificity.^54^ Collectively, these results suggest that two types of oncogenic mechanisms involving *KMT2A* fusions exist that may be marked with unique gene expression patterns.

We identified five *NPM1*-related clusters, further underpinning findings of transcriptional heterogeneity among *NPM1*-mutated patients^15,16,55^, but also providing additional insight into co-mutations and detailed subtypes. We observed several samples from *NPM1*-related clusters that lacked the *NPM1* mutation. Several rare *NPM1* fusions, like *NPM1::MLF1,* have been described to lead to cytoplasmic localisation of *NPM1*, comparable to the canonical *NPM1* frameshift.^45,56^ These non-canonical mishaps could lead to a *NPM1*-mutated-like presentation and similarities in survival and drug response should be explored. Additionally, NPM1(1)^T^ was mutually exclusive enriched for *IDH1/2* and *TET2* co-mutations. *IDH1/2* mutations lead to an aberrant alpha-ketoglutarate metabolism and are functionally complementary to TET2 loss-of-function mutations ^57^. While NPM1(1)^T^ and NPM1(5)^T^ show significant enrichments for *TET2*, only NPM1(1)^T^ shows this mutual exclusivity with *IDH1/2*. This suggests that only NPM1(1)^T^ is driven by aberrant alpha-ketoglutarate metabolism, which should be further studied using metabolomics.

Similarly, cytogenetic abnormalities, AML-MRC mutations and high MECOM expression were found in all AML-MRC clusters, but lead to different gene expression. A possible explanation could be clonal architecture and the differentiation state of the cell acquiring the leukemic aberration, both known to influence the biology of the resultant leukaemia.^58,59^ To our knowledge, we are the first to show different gene expression-based subgroups in AML-MRC, with divergent drug responses. Accurate identification of these clusters requires gene expression analysis, showing the relevance of our work.

We found no additional survival differences between other clusters. However, data availability limited the survival analysis, and different treatment protocols across studies could have led to confoundment. Survival differences between transcriptional subtypes should thus be further explored in one large cohort. However, we did find marked differences in drug responses between the clusters. Ideally, new studies should test in-patient efficacy of drugs with good ex-vivo responses in transcriptional subtypes. Furthermore, transcriptional subtyping could aid AML specialists in the highly complex field of clinical care and lead to multidisciplinary tailored-based treatment advice.^60^

In conclusion, the transcriptional subtypes reveal heterogeneity in AML not captured by genetic classification. Integration of transcriptomics into AML research and diagnostics could improve disease understanding and lead to more treatment options.

## Supporting information

Appendix 1, DEG

## Data Availability

The datasets generated and/or analysed during the current study are available from www.github.com/jeppeseverens/AMLmap. All code used to generate results is available on reasonable request. The predictor is available from www.github.com/jeppeseverens/AMLmapR as an R package.

https://www.github.com/jeppeseverens/AMLmap

https://www.ncbi.nlm.nih.gov/geo/query/acc.cgi?acc=GSE67040

https://ega-archive.org/dacs/EGAC00001000956

https://doi.org/10.1038/s41586-018-0623-z

https://www.vizome.org/aml

https://www.ocg.cancer.gov/programs/target/data-matrix

https://www.cbioportal.org/study/summary?id=laml_tcga_pub

https://data.leucegene.iric.ca/

https://portal.gdc.cancer.gov

## Acknowledgements

This project was funded by a strategic investment of the Leiden University Medical Center, embedded within the Leiden Oncology Center, and executed within the Leiden Center for Computational Oncology. EvdA was funded by a personal grant from the Dutch Research Council (NWO; VENI: 09150161810095). The funding bodies had no role in the study design, the collection, analysis, and interpretation of data, the writing of the manuscript, and the decision to submit the manuscript for publication.

## Author Contributions

M.J.R., M.G., and E.B.A. conceived and designed the project; E.B.A. acquired funding; E.B.A. performed project administration; M.G., E.B.A., H.V., R.R.B., C.J.M.H., P.B. performed oversight and management of resources (data generation, collection, transfer, infrastructure, data processing); J.F.S. performed computational and statistical analyses; J.F.S., E.B.A., M.G., E.O.K., E.S.-L. performed analyses and interpretation; J.F.S. performed and structured data visualisation; M.J.R., M.G. and E.B.A. provided supervision and scientific direction; J.F.S. wrote the manuscript; and all authors critically reviewed the manuscript and figures.

